# Free cortisol and free 21-deoxycortisol in the clinical evaluation of congenital adrenal hyperplasia

**DOI:** 10.1101/2024.07.11.24310065

**Authors:** Bas P.H. Adriaansen, Agustini Utari, André J. Olthaar, Rob C.B.M. van der Steen, Karijn J. Pijnenburg-Kleizen, Lizanne Berkenbosch, Paul N. Span, Fred C.G.J. Sweep, Hedi L. Claahsen-van der Grinten, Antonius E. van Herwaarden

**Author notes:** Corresponding author Bas P.H. Adriaansen, Radboud university medical Center, Department of Pediatrics, Geert Grooteplein Zuid 10 (huispost 804), Postbus 9101, 6500HB, Nijmegen, Gelderland, the Netherlands. These authors contributed equally to this work and share last authorship (Hedi L. Claahsen-van der Grinten, Antonius E. van Herwaarden). **Funding:** This work was funded by International Fund Congenital Adrenal Hyperplasia (IFCAH). **Disclosures:** The authors have nothing to disclose.

## Abstract

**Context:** Some patients with classic congenital adrenal hyperplasia (CAH) survive without glucocorticoid treatment. Increased precursor concentrations in these patients might lead to higher free (biological active) cortisol concentrations by influencing the cortisol-protein binding. In 21-hydroxylase deficiency (21OHD), the most common CAH form, accumulated 21-deoxycortisol (21DF), a precursor steroid, may further increase glucocorticoid activity. Both mechanisms could explain the low occurrence of symptoms in some untreated classic CAH patients.

**Objective:** Establishment and validation of an LC-MS/MS method for (free) cortisol and (free) 21DF to quantify these steroids in untreated patients with classic CAH (n=29), non-classic CAH (NCCAH, n=5), other forms of adrenal insufficiency (AI, n=3), and controls (n=11) before and 60 minutes after Synacthen® administration.

**Results:** Unstimulated total cortisol levels of untreated classic CAH patients (median 109 nmol/L) were lower compared to levels in untreated NCCAH patients (249 nmol/L, p=0.010) and controls (202 nmol/L, p=0.016), but free cortisol concentrations were similar. Basal free 21DF levels were high in 21OHD patients (median 5.32 nmol/L) and undetectable in AI patients and controls (<0.19 nmol/L). After Synacthen® administration, free concentrations of 21DF -but not cortisol-increased only in patients with 21OHD.

**Conclusions:** Free cortisol levels were similar in classic CAH compared to controls and NCCAH patients, suggesting a comparable availability of cortisol. Additionally, 21OHD patients produce high levels of the glucocorticoid 21DF, possibly explaining the low occurrence of symptoms in some classic 21OHD patients. Free cortisol and (free) 21DF levels should be considered in the clinical evaluation of adrenal insufficiency in patients with CAH.

## Introduction

Glucocorticoids, like cortisol, are among others essential for maintaining blood pressure, glucose metabolism, and modulating the immune response [1, 2]. Cortisol is produced by the adrenal cortex and has a diurnal rhythm with the highest concentration in the early morning [3]. The hypothalamus regulates this rhythm by stimulating the pituitary gland, which subsequently produces adrenocorticotropic hormone (ACTH). Negative feedback by cortisol balances the activity of the hypothalamus-pituitary-adrenal (HPA) axis. During periods of physical stress, such as infection or surgery, the demand for cortisol increases, which is mediated by an increased ACTH release.

Patients with primary adrenal insufficiency (PAI) have insufficient cortisol production and are unable to increase cortisol levels during periods of stress. This leads to symptoms of cortisol deficiency such as extreme fatigue, weight loss, and abdominal pain. In adults, PAI is most often caused by acquired conditions such as auto-immune adrenalitis, known as Addison’s disease. In children, PAI is generally caused by inherited conditions affecting adrenal steroid production. The most common inherited form of PAI is classic congenital adrenal hyperplasia (CAH) caused by an enzyme deficiency in the adrenal steroidogenesis pathway. Most CAH patients have 21-hydroxylase deficiency (21OHD) (90 – 95%) or 11-hydroxylase deficiency (11-OHD) (∼5%). These patients have insufficient cortisol production and consequently, a reduced negative feedback to the pituitary gland, causing chronically elevated ACTH levels. This leads to continuous activation of the adrenal cortex and accumulation of precursor steroids before the enzymatic block. These precursor steroids are partly shunted into the unaffected adrenal androgen pathway, leading to hyperandrogenism. So, in contrast to other forms of PAI, the hallmark of classic CAH is not only a decreased concentration of cortisol but also strongly elevated concentrations of precursor steroids and adrenal androgens. Patients with non-classic CAH (NCCAH), a less severe subgroup, generally produce normal concentrations of cortisol but still have elevated concentrations of precursor steroids and androgens [4].

The first step in diagnosing PAI is measuring morning total cortisol concentrations in blood. To evaluate the potential responsiveness of the HPA axis to stress, stimulation tests are performed, such as the insulin tolerance test (ITT) [5] or the Synacthen® test [6]. In these tests, a suboptimal cortisol response is defined as a total cortisol concentration <500 nmol/L (18 μg/dL) [7], but there is an ongoing debate on this threshold [8]. In the CAH guideline, the cut-off is defined as <400 – 500 nmol/L (<14 – 18 μg/dL), depending on the measuring method [9].

Cortisol is in blood mostly bound to proteins (±95%), such as corticosteroid-binding globulin (CBG) and albumin [10, 11]. The measured total cortisol is comprised of the bound and free (i.e., unbound) cortisol, while, according to the free hormone hypothesis, only the latter determines the biological glucocorticoid activity [12]. In addition, patients with 21OHD have elevated levels of 17-hydroxyprogesterone (17OHP) that can be metabolized by 11-hydroxylase into 21-deoxycortisol (21DF) [13]. Interestingly, previous studies have described untreated classic CAH patients with insufficient cortisol production without overt complaints of cortisol deficiency [14–17]. Our group showed that 21DF can transactivate the glucocorticoid receptor with a relative potency of 49% compared to cortisol, the most potent endogenous glucocorticoid. Hence, 21DF contributes to the glucocorticoid activity, especially when the concentration is significantly increased, such as in patients with 21OHD [14]. In addition, it is known that precursor steroids can bind to CBG [11, 18, 19], thereby competing with cortisol for bindings spots on CBG and preventing cortisol from binding, therewith increasing the free fraction of cortisol. These mechanisms might contribute to the attenuation of symptoms in patients with untreated classic CAH. So far, no studies have been conducted on free cortisol and free 21DF measurements in patients with CAH. Our aim is to quantify total and free concentrations of cortisol and 21DF in the morning and 60 minutes after administration of Synacthen® in patients with untreated classic CAH (both 11OHD and 21OHD). We will compare these results with patients with NCCAH, patients with adrenal insufficiency (AI) but not CAH, and controls to investigate the situation of CAH patients in more detail. We hypothesize that free cortisol and (free) 21DF concentrations might give a better reflection of the glucocorticoid activity in patients with 21OHD compared to solely measuring total cortisol levels.

## Materials and methods

### Subjects

This study was approved by the local medical ethics committee (Radboudumc, case number 2021-12944). Patients were included via three routes (see Figure S1):

(I) Indonesian cohort (n=22): Serum samples from an Indonesian cohort of untreated classic CAH patients (21OHD, n=17; 11OHD, n=5) were obtained as previously described [14]. Ethical approval was obtained from the ethical committee of the Faculty of Medicine, Diponegoro University, Semarang, Indonesia (713/EC/FK-RSDK/2016).
(II) Dutch retrospective cohort (n=39): Stored residual serum samples from Synacthen® tests performed between January 2016 and March 2021 were retrospectively collected.
(III) Dutch prospective cohort (n=19): Residual serum and plasma samples were obtained from three hospitals in the Netherlands between April 2021 and January 2024.

For the Synacthen® tests, blood was obtained before (T_0_) and 60 minutes after (T_60_) administration of 250 μg Tetracosactide. Samples were kept frozen at ≤ −30 °C until analysis. Inclusion criteria were: ≥300 µL residual serum available and patients not treated with systemic glucocorticoids for at least one year before blood withdrawal. Pseudonymized data about age, gender, time of blood withdrawal, indication of Synacthen® test, and diagnosis were collected. Patients were divided into four groups: (I) patients with classic CAH (both 21OHD and 11OHD), (II) patients with NCCAH based on 21OHD, (III) patients with adrenal insufficiency (AI) but not CAH, and (IV) patients with an optimal cortisol response after Synacthen® (controls). Classification of CAH patients in classic CAH and NCCAH was based on clinical characteristics, biochemical evaluation, and genetic analysis (Table S1) [20, 21]. Sequencing results were compared to the reference sequence GenBank NM_000500.9 for *CYP21A2* and NM_000497.3 for *CYP11B1*. In this article the term ‘classic CAH’ refers to both 21OHD and 11OHD patients.

### Measurement of total steroid concentrations with LC-MS/MS

Total serum 17OHP (17-hydroxypregn-4-ene-3,20-dione), 11-deoxycortisol (17,21-dihydroxypregn-4-ene-3,20-dione), 21DF (11,17-dihydroxypregn-4-ene-3,20-dione), and cortisol (11,17,21-trihydroxypregn-4-ene-3,20-dione) were analyzed by a clinically validated liquid chromatography tandem mass spectrometry (LC-MS/MS) method on a 6490 triple quadrupole LC-MS/MS (Agilent Technologies, Santa Clara, CA) preceded by protein precipitation and solid phase extraction (SPE). An internal standard mix (20 µL in methanol:H_2_O 30:70) containing [^13^C_3_]-17OHP, [^2^H_5_]-11-deoxycortisol, [^2^H_4_]-21DF, and [^13^C_3_]-cortisol (all from Isosciences, Ambler, PA) with concentrations of 10, 10, 20, and 100 nmol/L, respectively, was added to 100 µL serum. For protein precipitation, acetonitrile + 0.1% formic acid was added and incubated for at least 10 minutes at room temperature and centrifuged (10 min, 5000g). Then, 200 μL supernatant was added to 300 μL H_2_O followed by SPE using Oasis® HLB 1cc cartridges (Waters Corporation, Milford, MA). These cartridges were preequilibrated with 1 mL methanol:isopropanol (95:5) and consequently washed with 1 mL H_2_O. After sample application, columns were washed with 1 mL H_2_O and 1 mL methanol:H_2_O (30:70). The eluate (300 μL, methanol:isopropanol 95:5) was dried under a stream of N_2_ gas at 40 °C, reconstituted in 100 μL methanol:H_2_O (30:70) and injected (10 µL) into an Agilent Technologies 1290 Infinity UHPLC-system (Agilent Technologies). LC-MS/MS settings are shown in Table S2. The collision energy was optimized for each analyte and internal standard and two mass transitions (quantitative and qualitative) were monitored. A nine-point calibration curve with 17OHP, 11DF, 21DF, and cortisol (all from Sigma-Aldrich, St. Louis, MO, USA) was used for quantification. The calibration ranges were 0 – 167 nmol/L for 17OHP, 0 – 191 nmol/L for 11DF, 0 – 167 nmol/L for 21DF, and 0 – 2,299 nmol/L for cortisol. In-house quality controls of human serum were measured in duplicate in each run.

### Measurement of free steroid concentrations with LC-MS/MS

Measurement of free 21DF was incorporated in a clinically validated measuring method for free cortisol and free testosterone. Equilibrium dialysis was performed with samples buffered to pH 7.4 (± 0.03) at 37 °C (± 0.5) by a previously described HEPES buffer (4-(2-Hydroxyethyl)piperazine-1-ethanesulfonic acid; Merck, Rahway, NJ, USA) that resembled the ionic environment of serum [22]. Two identical volume compartments (180 μL) were separated by a semipermeable regenerated cellulose membrane (Serva Electrophoresis GmbH, Heidelberg, Germany) with a diameter of 28 mm and maximum permeability of 5 kDA. During dialysis, cells were placed in a water bath at 37 °C (± 0.5) for 5h while continuously rotating. After dialysis, dialysate was emptied from the cell and SPE was performed with 100 μL preceded by the addition of H_2_O (30 μL) and an internal standard mix (20 μL) containing [^2^H_4_]-21DF, [^13^C_3_]-cortisol, and [^13^C_3_]-testosterone (all from IsoSciences). For SPE, Oasis® MCX 1cc cartridges (Waters Corporation) were preequilibrated with 1 mL methanol:isopropanol (95:5) and consequently washed with 1 mL H_2_O. After sample application, columns were washed with 1 mL H_2_O + 5% ammonium hydroxide and 1 mL methanol:H_2_O (20:80) + 2% formic acid. The eluate (300 μL, 100% methanol) was dried in the Savant™ SpeedVac™ Concentrator (ThermoFisher Scientific, Waltham, MA, USA) for ± 1h, reconstituted in 30 μL methanol:H_2_O (30:70) and injected (10 µL) into a Xavi® TQ-XS StepWave XS^TM^ triple quadrupole spectrometer (Waters Incorporation). LC-MS/MS settings including collision energy and mass transitions are shown in Table S3. A nine-point calibration curve with 21DB (Steraloids, Newport, RI, USA), 21DF, and cortisol (both from Sigma-Aldrich) was used for quantification. The calibration ranges were 0 – 115 nmol/L for all three steroids.

### Method validation

All measurements were performed in an ISO15189 accredited laboratory. All clinically validated measuring methods are monitored for quality in each run using in duplicate measurements of in-house quality controls of human serum. For validation, a CLSI EP6 protocol [23] in serum was used to assess linearity. We measured steroid concentrations in a blank sample that was injected after the highest standard to assess carry-over. Ion suppression was measured according to CLSI C62-A guideline [24]: qualitative ion suppression by continuous infusion of the labeled steroid, and quantitative ion suppression by comparing the signal of human serum measurements (n=5) with the signal of spiked prepared matrix-free blank. Imprecision was assessed by a modified CLSI EP5 protocol [25] with pooled human serum samples at a low (n=7 for 21DF; n=20 for cortisol) and high concentration (n=8 for 21DF, n=20 for cortisol). The lower limit of quantification (LLOQ) was defined as the lowest concentration with an inter-assay coefficient of variation (CV) <20% and was determined with a modified EP evaluator protocol from pooled serum by between-day assay repeated measurements (n=10). For 21DF, recovery was calculated in five samples to which 29.94 nmol/L was added. For cortisol, a National Institute of Standards and Technology (NIST) standard (LOT number: 271020) diluted to 1.496 nmol/L was measured each run (n=29). A correction factor of 1.03 was applied to adjust free cortisol levels according to the NIST standard. Possible interfering substances were identified by measuring commercially available steroids and anti-androgens with a molecular mass ≤2 g/mol lower than the analyte or the internal standard. First, retention time interference was checked, and second, MRM transition interference was evaluated.

### Data and statistical analyses

Free cortisol percentage was calculated by dividing the free cortisol concentration by the total cortisol concentration from the same sample multiplied by 100%. Since cortisol has a diurnal rhythm, only morning samples (between 08.00 – 11.00 AM) were used for the analyses of absolute free and total cortisol concentrations. Results <LLOQ with a signal-to-noise ratio <5 were assigned a value of half the LLOQ. These results were excluded from the calculation of percentages. A Kruskal-Wallis test was used to compare means between different subgroups (classic CAH vs NCCAH vs AI vs control). For comparison of related samples (T_0_ vs T_60_), a related-samples Wilcoxon signed ranked test was used.

## Results

### Subjects

In total, data from 22 Indonesian and 58 Dutch patients were collected, see Figure S1 for the inclusion flowchart. Patients with an insufficient volume of residual material (n=31) and patients treated with glucocorticoids before the test were excluded (n=1). In total, 48 patients were divided into four groups: classic CAH (n=29; 21OHD [n=24] and 11OHD [n=5]), NCCAH (n=5), AI (n=3), and controls (n=11). Demographics of included patients are shown in Table 1.

**Table 1.** Demographics of included patients. Continuous data is given as median (IQR).

|  | CAH | NCCAH | AI | Control |
| --- | --- | --- | --- | --- |
| Number (n) | 29 | 5 | 3 | 11 |
| Age (years) | 13.2<br>(3.3 – 19.6) | 8.9<br>(6.0 – 18.7) | 16.1<br>(11.8 – 17.3) | 9.8<br>(7.9 – 14.7) |
| Genotype<br>(% 46,XY) | 10.3 | 40.0 | 33.3 | 63.6 |
| Sex (% males) | 41.4 | 40.0 | 33.3 | 63.6 |
| Diagnosis /<br>Indication for<br>Synacthen® test | CAH<br>(100%) | NCCAH<br>(100%) | X-ALD (33.3%)<br>AD (33.3%)<br>Unknown<br>(33.3%) | Screening X-ALD (18%)<br>Premature pubarche (18%)<br>Screening hypothyroidism (18%)<br>Fatigue (9%)<br>Collapse and hypotension (9%)<br>Screening MEN-1 (9%)<br>Rapid postnatal growth (9%)<br>Once measured low cortisol (9%) |

### Method validation

An LC-MS/MS method for free cortisol and free 21DF was established and validated. The measurement of 21DF was linear between 0.13 and 99.2 nmol/L and the measurement of free cortisol was linear between 0.08 and 66 nmol/L. The maximum deviation from linearity, imprecision, LLOQ, recovery, and ion suppression are shown in Table 2. Carry-over was negligible for both components. For cortisol and its internal standard, we evaluated 18-hydroxycorticosterone as possible interfering substance and it showed no interference. For 21DF and its internal standard, corticosterone, 11-deoxycortisol, and 18-hydroxy-11-deoxycorticosterone were evaluated and showed no interference.

**Table 2.** Linearity, Imprecision (within and between day) on low and high level, lower limit of quantification (LLOQ), recovery, and ion suppression for free 21DF and free cortisol measured by LC-MS/MS after dialysis.

|  | **Free 21DF** | **Free Cortisol** |
| --- | --- | --- |
| **Linearity** |  |  |
| *Linearity range (nmol/L)* | 0.13 – 99.2 | 0.08 – 66.0 |
| *Deviation from linearity (%)* | 1.55 | 0.96 |
| **Imprecision low** |  |  |
| *Concentration (nmol/L)* | 0.36 | 11.4 |
| *Within day (%)* | 2.8 | 2.6 |
| *Between day (%)* | 8.5 | 4.6 |
| **Imprecision high** |  |  |
| *Concentration (nmol/L)* | 4.89 | 104.4 |
| *Within day (%)* | 3.3 | 2.5 |
| *Between day (%)* | 2.8 | 4.0 |
| **LLOQ (nmol/L)** | 0.19 | <0.32* |
| **Recovery** |  |  |
| *Concentration (nmol/L)* | 29.94 | 1.496 |
| *Average recovery (%)* | 108 | 97.3 |
| **Ion suppression (%)** | <1 | <1 |

\*mean CV of 6.1%

#### Baseline

##### Morning free cortisol levels in classic CAH patients are similar to levels in controls at T _0_

The median total cortisol concentration in the morning (samples at T_0_) was 106 nmol/L in the untreated classic CAH patients, which was significantly lower than NCCAH patients (249 nmol/L, p=0.010) and controls (202 nmol/L, p=0.016) (Figure 1A).

**Figure 1.**
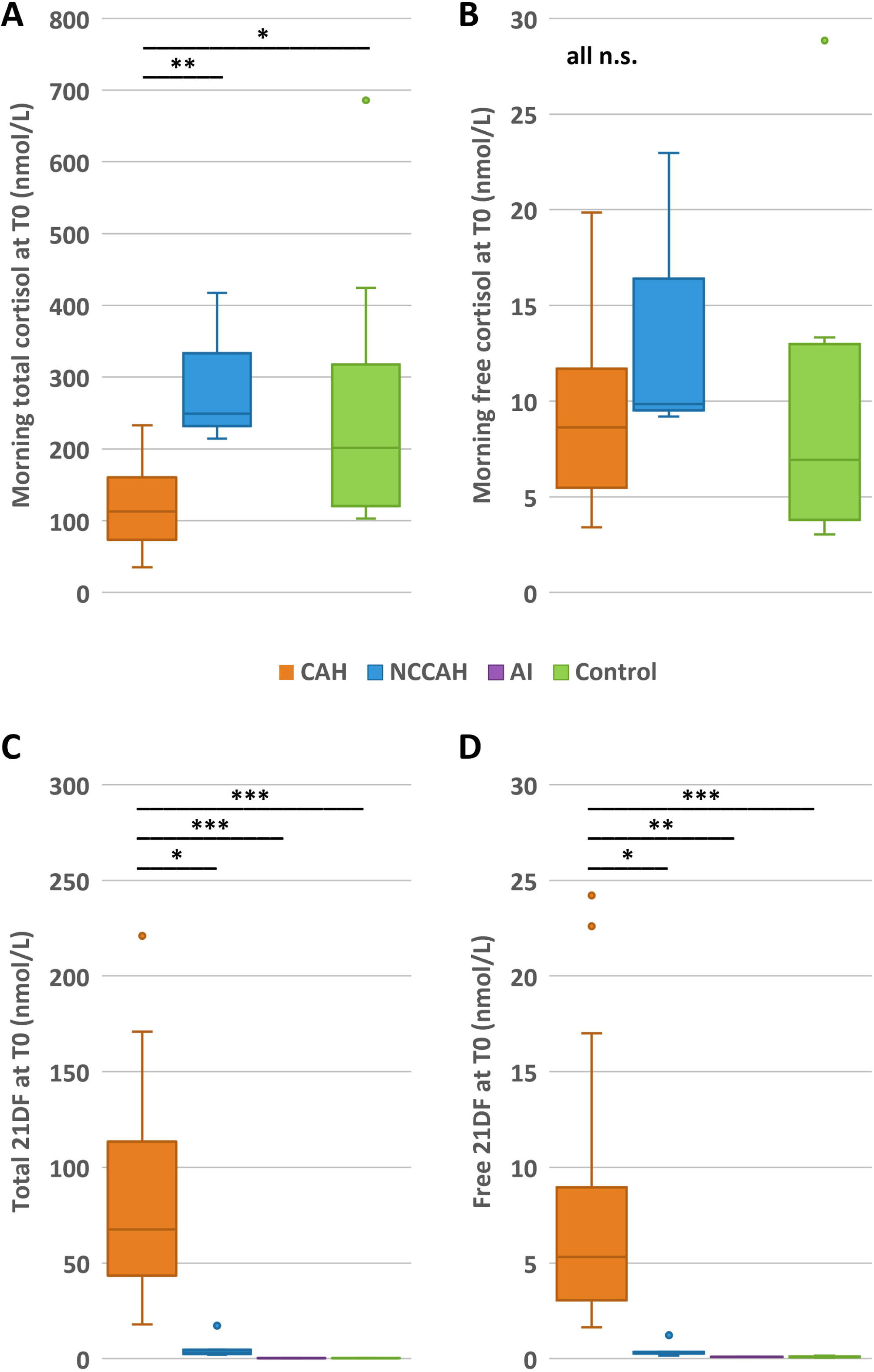
Total and free cortisol and 21DF concentrations (nmol/L) at T0 in patients with classic congenital adrenal hyperplasia (CAH), non-classic congenital adrenal hyperplasia (NCCAH), adrenal insufficiency but not CAH (AI), and controls. (A) Morning (8.00 – 11.00 AM) concentrations of total cortisol; (B) Morning concentrations of free cortisol; (C) total 21DF concentrations (D) Free 21DF concentrations. n.s. non-significant, *p≤0.05, ** p≤0.01, ***p≤0.001.

Interestingly, the median *free* morning cortisol concentration in untreated classic CAH patients (8.62 nmol/L) was not significantly different from NCCAH patients (9.84 nmol/L, p=0.313) and controls (6.94 nmol/L, p=0.573) (Figure 1B). So, in contrary to the total cortisol concentrations, this suggests a comparable availability of cortisol in our classic CAH cohort compared to NCCAH patients and controls.

In untreated classic CAH patients, the concentration of free cortisol is higher than expected based on the total cortisol concentration. This is also indicated by the percentage of free cortisol, which was 7.5% for classic CAH patients, significantly higher than free cortisol percentages of NCCAH patients (5.5%, p=0.025), AI patients (2.1%, p<0.001), and controls (3.3%, p<0.001) (Figure S2A).

##### Patients with 21OHD have increased total 21DF concentrations compared to AI patients and controls

Patients with 21OHD have increased total 21DF concentrations with a median of 67.5 nmol/L at T_0_. This was significantly higher compared to all other groups: NCCAH patients had a median of 3.03 nmol/L (p=0.020), and total 21DF concentrations in AI patients and controls were all <LLOQ (0.7 nmol/L) (p<0.001) (Figure 1C).

For *free* 21DF concentrations at T_0_, the same is observed: the median concentration in 21OHD patients (5.32 nmol/L) was significantly higher compared to median levels in NCCAH patients (0.34 nmol/L; p=0.025), AI patients (<0.19 nmol/L, (p=0.003), and controls (<0.19 nmol/L, p<0.001) (Figure 1D).

In all 11OHD patients, total and free 21DF concentrations were <LLOQ (data not shown).

#### After Synacthen® stimulation

##### Patients with classic CAH have lower (free) cortisol concentrations T_60_ compared to controls

All controls had a total cortisol concentration ≥500 nmol/L at T_60_ (median 658 nmol/L), indicating sufficient cortisol production during stress according to current guidelines [7]. In NCCAH patients, total cortisol concentrations increased with a median of 42% (p=0.043), although not with the same steep increase as observed in controls (198%; p=0.005). In none of the NCCAH patients the 500 nmol/L threshold (median 402 nmol/L) was reached, but 60% (3/5) of them reached the 400 nmol/L threshold as proposed in the CAH guideline [9]. In the remaining two NCCAH patients, stimulated total cortisol concentrations of 305 and 397 nmol/L were measured. Total cortisol concentrations did not increase upon Synacthen® administration in classic CAH and AI patients (p=0.466 and p=0.655, respectively) (Figure 2A). Classic CAH (median 119 nmol/L) and AI patients (median 172 nmol/L) did both not reach the 500 nmol/L threshold [7] nor the 400 nmol/L threshold [9], indicating adrenal insufficiency, as expected in these patients.

**Figure 2.**
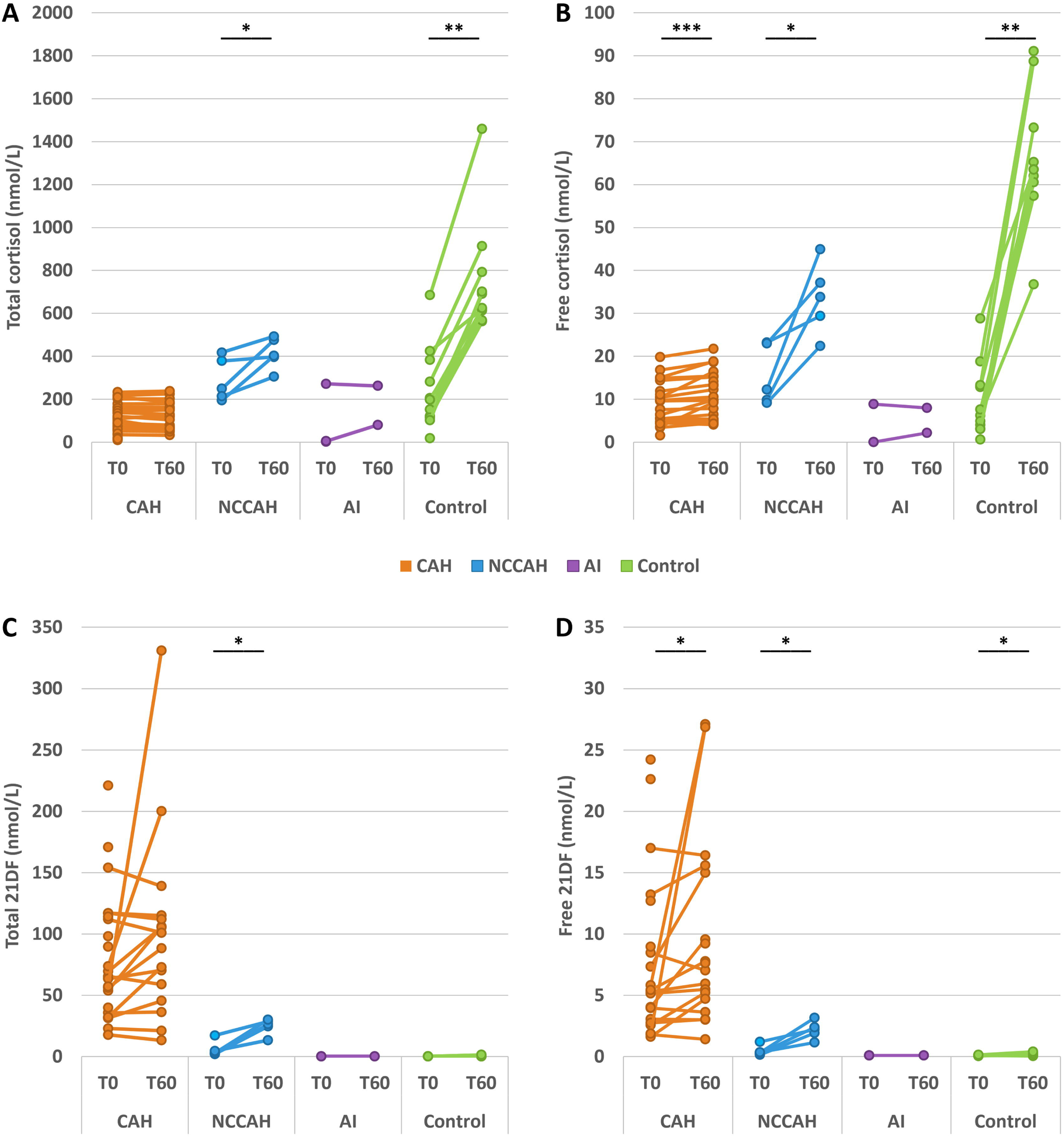
Increase of total and free cortisol and 21DF concentrations (nmol/L) before (T0) and after (T60) Synacthen® in patients with classic congenital adrenal hyperplasia (CAH), non-classic congenital adrenal hyperplasia (NCCAH), adrenal insufficiency but not CAH (AI), and controls. (A) Change in total cortisol concentrations; (B) Change in free cortisol concentrations; (C) Change in total 21DF concentrations; (D) Change in free 21DF concentrations. *p≤0.05, ** p≤0.01, ***p≤0.001.

The *free* cortisol concentration increased after Synacthen® administration in classic CAH patients by 10.7% (p=0.001). In NCCAH patients and controls, we observed a median increase of 144% (p=0.043) and 610% (p=0.005), respectively. No significant increase in the free cortisol concentration was observed in AI patients (p=0.655) (Figure 2B). At T_60_, the median *free* cortisol concentration in classic CAH patients was 9.85 nmol/L, which was significantly lower compared to levels in NCCAH patients (33.8 nmol/L, p=0.010) and controls (63.6 nmol/L, p<0.001), but not significantly different from AI patients (5.08 nmol/L, p=0.377) (data not shown). In all controls, an increase of free cortisol ≥37 nmol/L after stimulation was measured. Levels of free cortisol after Synacthen® stimulation in NCCAH patients ranged from 22 to 45 nmol/L. Patients with classic CAH did not reach serum levels of free cortisol seen in controls after Synacthen® (maximum 21.8 nmol/L).

After Synacthen®, the *percentage of free cortisol* increased in classic CAH (p=0.001), NCCAH (p=0.043), and control patients (p=0.005), but not in AI patients (p=0.655) (Figure S2B). At T_60_, the median free cortisol percentage of classic CAH patients (8.9%) was significantly higher compared to AI patients (2.9%, p=0.019) but not significantly different from NCCAH patients (7.4%, p=0.235) and controls (9.7%, p=0.520) (Figure S2C).

##### Concentrations of free 21DF increase after Synacthen® administration in classic 21OHD patients

The total 21DF concentration significantly increased upon ACTH stimulation in NCCAH patients (p=0.043), but not in classic 21OHD patients (p=0.121) (Figure 2C). At T_60_, the median total 21DF concentrations in classic 21OHD and NCCAH patients were 94.6 nmol/L and 26.4 nmol/L, respectively. The total 21DF concentration in AI patients remained <LLOQ at T_60_. In controls, the maximum total 21DF concentration was twice the LLOQ (0.7 nmol/L) for two patients. The 21OHD patients had significantly higher total 21DF concentrations at T_60_ compared to AI patients (p=0.005) and controls (p<0.001) (data not shown).

The free 21DF concentration significantly increased in both classic CAH (p=0.023) and NCCAH patients (p=0.043) (Figure 2D). At T_60_, the median free 21DF concentration was 7.34 nmol/L in classic CAH patients and 2.22 nmol/L in NCCAH patients (p=0.054). For all AI patients, the free 21DF concentration was <LLOQ at T_60_. Controls had a median of 0.18 nmol/L free 21DF at T_60_ (data not shown). In all 11OHD patients, total and free 21DF concentrations were <LLOQ (data not shown). In classic 21OHD patients, the percentage of free 21DF was 8.4% at T_0_ which significantly increased to 10.3% (p=0.035) at T_60_. For NCCAH patients, there was no significant increase in the percentage of free 21DF after ACTH stimulation, it was 8.8% both at T_0_ and T_60_ (data not shown).

## Discussion

In this study, we evaluated for the first time the concentrations of free cortisol and free 21DF in a group of untreated patients with classic CAH (both 21OHD and 11OHD), NCCAH, other forms of AI, and controls. We found that the *free* cortisol concentration in untreated classic CAH patients is similar to the concentration in controls in the unstimulated situation, despite decreased *total* cortisol levels. This could explain why some patients with classic 21OHD or 11OHD show no or only mild symptoms of cortisol deficiency in the basal situation. In addition, patients with classic 21OHD have elevated concentrations of total and free 21DF, which is not observed in AI patients and controls, increasing the glucocorticoid activity. After Synacthen®, classic CAH patients show no increase in their total cortisol levels and only a minimal increase in free cortisol levels, but we measured increased free levels of the glucocorticoid 21DF in 21OHD patients. This might explain why these patients survived periods of severe stress without glucocorticoid treatment.

For this study, we have developed an LC-MS/MS method to measure free cortisol and free 21DF. The mean free cortisol percentage in our control group (3.7%) was similar to the percentage found by others (3.95%) [26, 27]. We observed that the relative increase in free cortisol concentration (610%) after Synacthen® administration was higher compared to the relative increase in total cortisol (198%), which was also observed in other studies [26–28]. Therefore, evaluating free concentrations may be more informative than evaluating total concentrations.

Glucocorticoids exert a plethora of functions in the human body, and they are especially necessary during periods of stress. Therefore, two situations should be distinguished when interpreting glucocorticoid levels: the daily baseline situation and an episode with physical stress.

### Baseline situation

Cortisol in blood is mostly bound to CBG and albumin, but only its free form is biologically active according to the free hormone hypothesis [29]. We found that in the baseline situation, *total* cortisol concentrations of untreated classic CAH patients are lower compared to controls, but *free* cortisol concentrations were similar. This is supported by data from Waaijers *et al.,* that found cortisol concentrations within the reference range in hair of untreated classic CAH patients [30], suggesting similar long-term free cortisol levels between classic CAH patients and the reference population. This might be explained by several hypotheses: (I) In classic CAH patients, increased levels of structurally analogous steroids, such as 17OHP and 21DF, may disrupt or prevent cortisol from binding to CBG or albumin [11, 18, 19], leading to a higher percentage of free cortisol; (II) CAH patients have increased concentrations of adrenal androgens, which are known to decrease 11β-hydroxy steroid dehydrogenase type 2 activity, preventing the conversion of active cortisol into inactive cortisone [31]; (III) androgens are able to decrease CBG levels, thereby possibly increasing the free cortisol concentrations [32, 33].

In addition, we found that in the basal situation, classic 21OHD patients have increased 21DF concentrations, which confirms the findings of previous studies [34–36]. Our group showed that 21DF has a relative potency of 49% to activate the glucocorticoid receptor compared to cortisol [14]. The median free 21DF concentration in untreated classic CAH patients of 5.32 nmol/L would be equivalent to (0.49 × 5.32 =) 2.61 nmol/L cortisol. Hypothetically this adds to the glucocorticoid activity of classic 21OHD patients. So, these patients have more glucocorticoid activity than expected based on their total cortisol levels. This could explain why some classic CAH patients have lived without glucocorticoid medication during daily circumstances. Patients with 11OHD are not capable of producing 21DF, but their free cortisol levels were already similar to controls.

### During periods of stress

During periods of physical stress, such as illness or surgery, there is a higher demand for glucocorticoids. Classic CAH patients were not able to increase their total cortisol levels after ACTH stimulation. In our study untreated classic CAH patients did not reach the 400-500 nmol/L total cortisol threshold after Synacthen® administration [9]. We found only a small increase in free cortisol levels, but classic CAH patients did not reach the increase in free cortisol levels of the control group. Interestingly, we did observe an increase in the free concentration of the glucocorticoid 21DF in patients with 21OHD up to a median of 7.34 nmol/L. Given that 21DF is capable of activating the glucocorticoid receptor these increased concentrations may contribute significantly to the glucocorticoid activity. However, it is questionable if this concentration of free glucocorticoids is enough to reach sufficient glucocorticoid activity in these patients during periods of stress to prevent Addisonian crisis. Therefore, it is still not entirely clear how previously described untreated classic 21OHD patients have survived periods of physical stress, such as dengue fever and surgery, without glucocorticoid treatment [14–17]. Other protective measures might be important in these patients during stress episodes. Kroon *et al.* have reviewed that androgens can also contribute to the glucocorticoid activity either by direct binding to the glucocorticoid responsive elements or by assisted loading [31], in which androgen signal transduction opens the chromatin, enabling activated glucocorticoid receptors to bind the responsive elements and enhance glucocorticoid signaling. In addition, Hamrahian *et al.* showed that the percentage of free cortisol increases in healthy individuals after administration of synthetic ACTH [29]. The continuous ACTH pressure in classic CAH patients might also directly increase the free cortisol percentage, leading to more free cortisol and hence more glucocorticoid activity. It would be interesting to further investigate these hypotheses in the specific situations of patients with CAH.

### Indication for glucocorticoid treatment

Previously, we and others described classic CAH patients who have survived without glucocorticoid treatment despite low total cortisol levels and the inability to increase total cortisol levels during stress [14–17]. The similar concentration of free cortisol in untreated classic CAH patients compared to controls might explain this clinical observation. Intriguingly, in clinical practice, some classic CAH patients -mostly males and 46,XX individuals living as males-exhibit so few overt signs of adrenal cortex insufficiency that they refuse glucocorticoid medication [37, 38]. However, patients have to be counseled about the potential risks of being untreated; Besides the risk to develop an Addisonian crisis, elevated ACTH levels can lead to the development of testicular adrenal rest tumors and adrenal myelolipomas [37]. Furthermore, elevated levels of androgens and precursor steroids may contribute to an unfavorable cardiovascular risk profile [39].

Genotype-phenotype correlations in CAH are not always accurate. In our study, we also found a discrepancy between the expected phenotype based on genetics and the clinical phenotype, especially for the R356W mutation in the *CYP21A2* gene. Stoupa *et al.* [40] reported that 60% of their NCCAH patients had an inadequate cortisol response after Synacthen®, while none of them experienced periods of cortisol deficiency, highlighting that not all patients with an insufficient total cortisol concentrations have clinically relevant adrenal cortex insufficiency.

Our findings suggest that measuring total cortisol levels alone may not be the most appropriate way to assess adrenal cortex function in all forms of PAI. Supplementary measurement of free cortisol may provide a more accurate reflection of glucocorticoid activity. In 21OHD patients, the concentration of (free) 21DF should also be considered to comprehensively evaluate the glucocorticoid activity. This would be especially useful in patients with milder forms of CAH, to indicate the necessity of glucocorticoid treatment. More research is necessary to compute cut-off values for defining adrenal cortex insufficiency and to incorporate it in clinical practice. Measuring free cortisol levels is not only useful for CAH patients, but also for patients with other conditions in which there is a discrepancy between free and total cortisol levels, for instance patients with hypoproteinemia during critical illness [29], patients with CBG-deficiency [10], or patients who use oral contraceptive medication [41].

Our study has several strengths and limitations. This is the first study that quantified free cortisol and free 21DF concentrations in untreated classic CAH patients. We did not only compare levels with controls but also with patients with NCCAH and AI. Another strength is the use of a state-of-the-art dialysis method to obtain free concentrations followed by a sensitive LC-MS/MS method, which is the gold standard for steroid measurements in serum [9]. Furthermore, deuterium or C-isotopes of each steroid were used as internal standards to allow for the correction of matrix effects. Moreover, all included CAH patients were genetically confirmed. A limitation of our study is the small sample size of the study groups, especially AI patients. PAI is rare and the volume of residual serum was not always sufficient to perform the measurements. Cortisol has a diurnal rhythm, but not all samples were taken in the morning. To correct for this, we only included measurements before 11.00 am in the analyses for total and free cortisol.

To conclude, untreated classic CAH patients have free cortisol concentrations in the same range as controls under basal conditions. Moreover, patients with 21OHD have an increased production of total and free 21DF, a precursor steroid with glucocorticoid activity. Both mechanisms might explain the low occurrence or lack of symptoms of adrenal insufficiency in daily situations in some untreated classic CAH patients. During periods of stress, (free) 21DF concentrations increase in 21OHD patients, in contrast to patients with other forms of PAI, with probably additional protective effects. Future research should focus on the role of free cortisol and free precursor steroids, especially in milder forms of CAH to accurately assess the glucocorticoid activity and indicate the necessity of glucocorticoid treatment.

## Data Availability

All data produced in the present study are available upon reasonable request to the authors after acceptance

## Notes

### Competing Interest Statement

The authors have declared no competing interest.

### Author Declarations

The local medical ethics committee of the Radboud university medical center gave ethical approval for this work (case number 2021-12944).

