## Supplementary material for "Free cortisol and free 21-deoxycortisol in the clinical evaluation of congenital adrenal hyperplasia": Table S1; Table S2; Table S3; Figure S1; Figure S2

***Table S1*** *Genetic analyses of included CAH patients (n=34) with predicted phenotype based on residual enzyme activity and literature. Legacy names are given for pathogenic variants in CYP21A2 if possible, otherwise pathogenic variants and protein changes are given according to HGVS nomenclature rules (varnomen.hgvs.org). SW, salt wasting; SV, simple virilizing; NC non-classic.*

| **Group** | **Gene** | **Variant 1** | |  | **Variant 2** | | **Predicted phenotype based on genotype** | **Phenotype based on clinical and biochemical characteristics** |
| --- | --- | --- | --- | --- | --- | --- | --- | --- |
|  |  | **Pathogenic variant** | **Predicted enzymatic activity** |  | **Pathogenic variant** | **Predicted enzymatic activity** |  |  |
| Classic CAH patients | *CYP21A2* | deletion  exon 1-7 | 0% |  | deletion  exon 1-7 | 0% | SW | SW |
|  | *CYP21A2* | Delta8 bp | 0% |  | deletion  exon 1-3 | 0% | SW | SW |
|  | *CYP21A2* | I2G | 0% |  | I2G | 0% | SW | SV* |
|  | *CYP21A2* | I2G | 0% |  | I2G | 0% | SW | SV* |
|  | *CYP21A2* | R356W | 0% |  | R356W | 0% | SW | SW* |
|  | *CYP21A2* | R356W | 0% |  | R356W | 0% | SW | SV* |
|  | *CYP21A2* | R356W | 0% |  | R356W | 0% | SW | SV* |
|  | *CYP21A2* | R356W | 0% |  | deletion  exon 1-3 | 0% | SW | SW* |
|  | *CYP21A2* | R356W | 0% |  | c.586C>T / p.(Gln196*) | 0% | SW | SW* |
|  | *CYP21A2* | R356W | 0% |  | deletion  exon 1-3 | 0% | SW | SW |
|  | *CYP21A2* | R356W | 0% |  | c. 1155_157del / p. (Ile386del) | unknown | SW/SV | SW* |
|  | *CYP21A2* | deletion  exon 1-6 | 0% |  | no second mutation found | unknown | unkown | SV* |
|  | *CYP21A2* | I2G | 0-1% |  | R356W | 0% | SV/SW | SW* |
|  | *CYP21A2* | I2G | 0-1% |  | R356W | 0% | SV/SW | SW* |
|  | *CYP21A2* | I2G | 0-1% |  | deletion  exon 1-3 | 0% | SV/SW | SW |
|  | *CYP21A2* | I2G | 0-1% |  | I2G | 0-1% | SW/SV | SW |
|  | *CYP21A2* | I172N | 1-5% |  | W19X | 0% | SV | SV* |
|  | *CYP21A2* | I172N | 1-5% |  | deletion  exon 1-6 | 0% | SV | SV* |
|  | *CYP21A2* | I172N | 1-5% |  | R356W | 0% | SV | SV* |
|  | *CYP21A2* | I172N | 1-5% |  | R483P | 0-1% | SV | SV |
|  | *CYP21A2* | I172N | 1-5% |  | I2G | 0-1% | SV | SV |
|  | *CYP21A2* | I172N | 1-5% |  | I172N | 1-5% | SV | SV* |
|  | *CYP21A2* | I172N | 1-5% |  | I172N | 1-5% | SV | SV* |
|  | *CYP21A2* | P30L | 20-30% |  | P30L | 20-30% | NC | SV* |
|  | *CYP11B1* | c. 799G>A / p.(Val252fs) | 0% |  | c. 799G>A / p.(Val252fs) | 0% | Classic | Classic |
|  | *CYP11B1* | c. 799G>A / p.(Val252fs) | 0% |  | c. 799G>A / p.(Val252fs) | 0% | Classic | Classic |
|  | *CYP11B1* | c. 799G>A / p.(Val252fs) | 0% |  | c. 799G>A / p.(Val252fs) | 0% | Classic | Classic |
|  | *CYP11B1* | c. 799G>A / p.(Val252fs) | 0% |  | c. 799G>A / p.(Val252fs) | 0% | Classic | Classic |
|  | *CYP11B1* | c. 799G>A / p.(Val252fs) | 0% |  | c. 799G>A / p.(Val252fs) | 0% | Classic | Classic |
| Non-classic CAH patients | *CYP21A2* | P453S | 30-50% |  | deletion | 0% | NC | NC |
|  | *CYP21A2* | V281L | 30-50% |  | R356W | 0% | NC | NC |
|  | *CYP21A2* | V281L | 30-50% |  | I172N | 1-5% | NC | NC |
|  | *CYP21A2* | V281L | 30-50% |  | I172N | 1-5% | NC | NC |
|  | *CYP21A2* | V281L | 30-50% |  | V281L | 30-50% | NC | NC |

**previously described in Engels et al. (2019) [5]*

***Table S2*** *LC-MS/MS settings total serum concentrations. Mobile phase A,* *methanol:H_2_O 20:80 + 2 mM NH_4_CH_3_COO + 0.1% formic acid; Mobile phase B, methanol:H_2_O 98:2 + 2 mM NH_4_CH_3_COO + 0.1% formic acid. RT, retention time; MRM, multiple reaction monitoring; CE, collision energy.*

| **Liquid chromatography settings** | | | | | | | | |
| --- | --- | --- | --- | --- | --- | --- | --- | --- |
| System | | 1290 Infinity UHPLC-system (Agilent Technologies) | | | | | | |
| Analytical column | | BEH C18 (50 x 2.1 mm, 1.7 µm) | | | | | | |
| Column temperature | | 60°C | | | | | | |
| Flow | | 0.4 mL/min | | | | | | |
| Total run time | | 8.0 minutes | | | | | | |
| **Time (min)** | | **A (%)** | **B (%)** | |  |  |  |  |
| 0.0 | | 70 | 30 | |  |  |  |  |
| 2.5 | | 70 | 30 | |  |  |  |  |
| 6.0 | | 40 | 60 | |  |  |  |  |
| 6.5 | | 2 | 98 | |  |  |  |  |
| 7.0 | | 2 | 98 | |  |  |  |  |
| 7.5 | | 70 | 30 | |  |  |  |  |
| 8.0 | | 70 | 30 | |  |  |  |  |
| **Mass spectrometry settings** | | | | | | | | |
| System | | 6490 triple quadrupole mass spectrometer (Agilent Technologies) | | | | | | |
| Ionisation mode | | Positive electron spray | | | | | | |
| Capillary Voltage | | 3.5 kV | | | | | | |
| Nebulizer | | 40 psi | | | | | | |
| Source gas temperature | | 150°C | | | | | | |
| Source gas flow | | 11 L/min | | | | | | |
| Sheath Gas temperature | | 350°C | | | | | | |
| Sheath Gas flow | | 12 L/min | | | | | | |
| Fragmentor voltage | | 380 V | | | | | | |
| **Steroid or**  **internal standard** | **RT (min)** | **Quantifier** | | |  | **Qualifier** | | **Dwell time (ms**) |
|  |  | **MRM** | | **CE (eV)** |  | **MRM** | **CE (eV)** |  |
| 17OHP | 4.70 | 331.3 > 97.1 | | 31 |  | 331.3 > 109.1 | 31 | 60 |
| [^13^C_3_]-17OHP | 4.70 | 334.3 > 100.1 | | 30 |  | 334.3 > 112.1 | 33 | 60 |
| 11DF | 2.58 | 347.2 > 97.1 | | 29 |  | 347.2 > 109.1 | 31 | 40 |
| [^2^H_5_]-11DF | 2.54 | 352.3 > 100.1 | | 31 |  | 352.3 > 113.1 | 29 | 40 |
| 21DF | 2.05 | 347.2 > 269.0 | | 18 |  | 347.2 > 97.0 | 33 | 40 |
| [^2^H_4_]-21DF | 2.05 | 351.2 > 273.0 | | 18 |  | 351.2 > 97.0 | 33 | 40 |
| cortisol | 1.45 | 363.4 > 121.1 | | 25 |  | 363.4 > 97.1 | 34 | 100 |
| [^13^C_3_]-cortisol | 1.45 | 366.4 > 124.1 | | 25 |  | 366.4 > 100.1 | 35 | 100 |

***Table S3*** *LC-MS/MS settings free serum concentrations. Mobile phase A,* *methanol:H_2_O 20:80 + 2 mM NH_4_CH_3_COO + 0.1% formic acid; Mobile phase B, methanol:H_2_O 98:2 + 2 mM NH_4_CH_3_COO + 0.1% formic acid. RT, retention time; MRM, multiple reaction monitoring; CE, collision energy.*

| **Liquid chromatography settings** | | | | | | | | |
| --- | --- | --- | --- | --- | --- | --- | --- | --- |
| System | | Acquity two-dimensional UPLC-system (Waters Corporation) | | | | | | |
| Analytical column | | BEH C18 (50 x 2.1 mm, 1.7 µm) | | | | | | |
| Column temperature | | 60°C | | | | | | |
| Flow | | 0.4 mL/min | | | | | | |
| Total run time | | 9.0 minutes | | | | | | |
| **Time (min)** | | **A (%)** | **B (%)** | |  |  |  |  |
| 0.0 | | 70 | 30 | |  |  |  |  |
| 5.0 | | 70 | 30 | |  |  |  |  |
| 5.5 | | 2 | 98 | |  |  |  |  |
| 6.5 | | 2 | 98 | |  |  |  |  |
| 7.0 | | 70 | 30 | |  |  |  |  |
| 9.0 | | 70 | 30 | |  |  |  |  |
| **Mass spectrometry settings** | | | | | | | | |
| System | | Xevo® TQ-XS StepWave XS^TM^ triple quadrupole mass spectrometer (Waters Corporation) | | | | | | |
| Ionisation mode | | Positive electron spray | | | | | | |
| Capillary Voltage | | 1.0 kV | | | | | | |
| Cone Voltage | | 20 V | | | | | | |
| Source temperature | | 150°C | | | | | | |
| Desolvation temperature | | 600°C | | | | | | |
| Desolvation gas flow | | 1,200 L/hr (N_2_) | | | | | | |
| Cone gas flow | | 150 L/hr | | | | | | |
| Collision gas flow | | 0.15 mL/min (Argon) | | | | | | |
| **Steroid or**  **internal standard** | **RT (min)** | **Quantifier** | | |  | **Qualifier** | | **Dwell time (ms**) |
|  |  | **MRM** | | **CE (eV)** |  | **MRM** | **CE (eV)** |  |
| 21DB | 4.41 | 331.2 > 295.2 | | 15 |  | 331.2 > 121.1 | 20 | 121 |
| [^13^C_3_]-T | 4.38 | 292.2 > 100.1 | | 20 |  | 292.2 > 112.1 | 24 | 121 |
| 21DF | 2.03 | 347.2 > 311.3 | | 15 |  | 347.2 > 269.3 | 17 | 121 |
|  |  |  |  |  |  | 347.2 > 121.2 | 25 | 121 |
| [^2^H_4_]-21DF | 2.03 | 351.2 > 315.3 | | 15 |  | 351.2 > 273.3 | 17 | 121 |
|  |  |  |  |  |  | 351.2 > 121.2 | 25 | 121 |
| cortisol | 1.46 | 363.2 > 121.1 | | 23 |  | 363.2 > 97.1 | 29 | 121 |
| [^13^C_3_]-cortisol | 1.45 | 366.2 > 124.2 | | 23 |  | 366.2 > 100.1 | 29 | 121 |

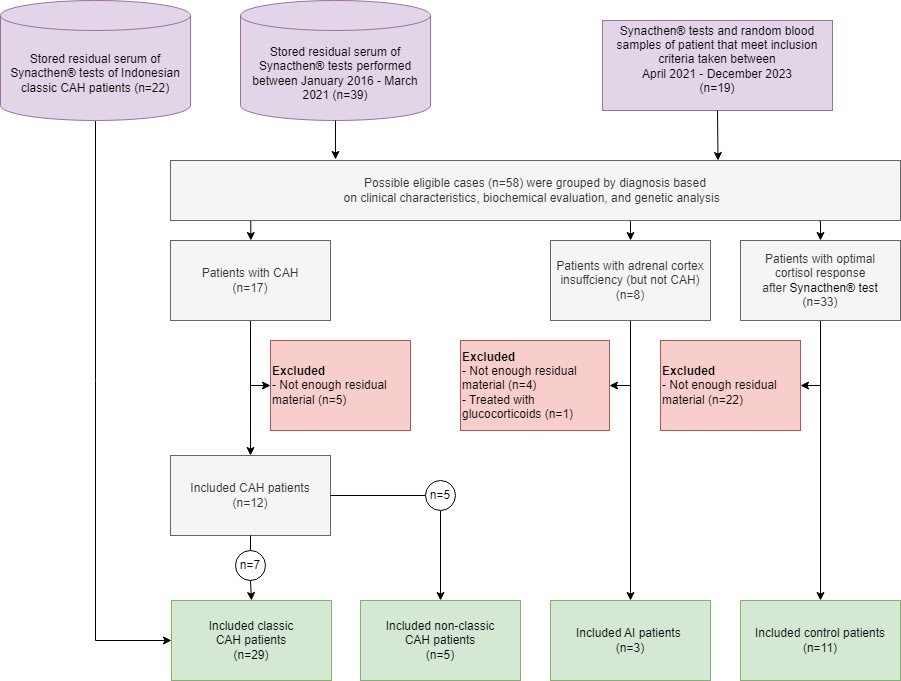

***Figure S1*** *Flowchart of included patients. CAH, congenital adrenal hyperplasia; AI, adrenal insufficiency.*

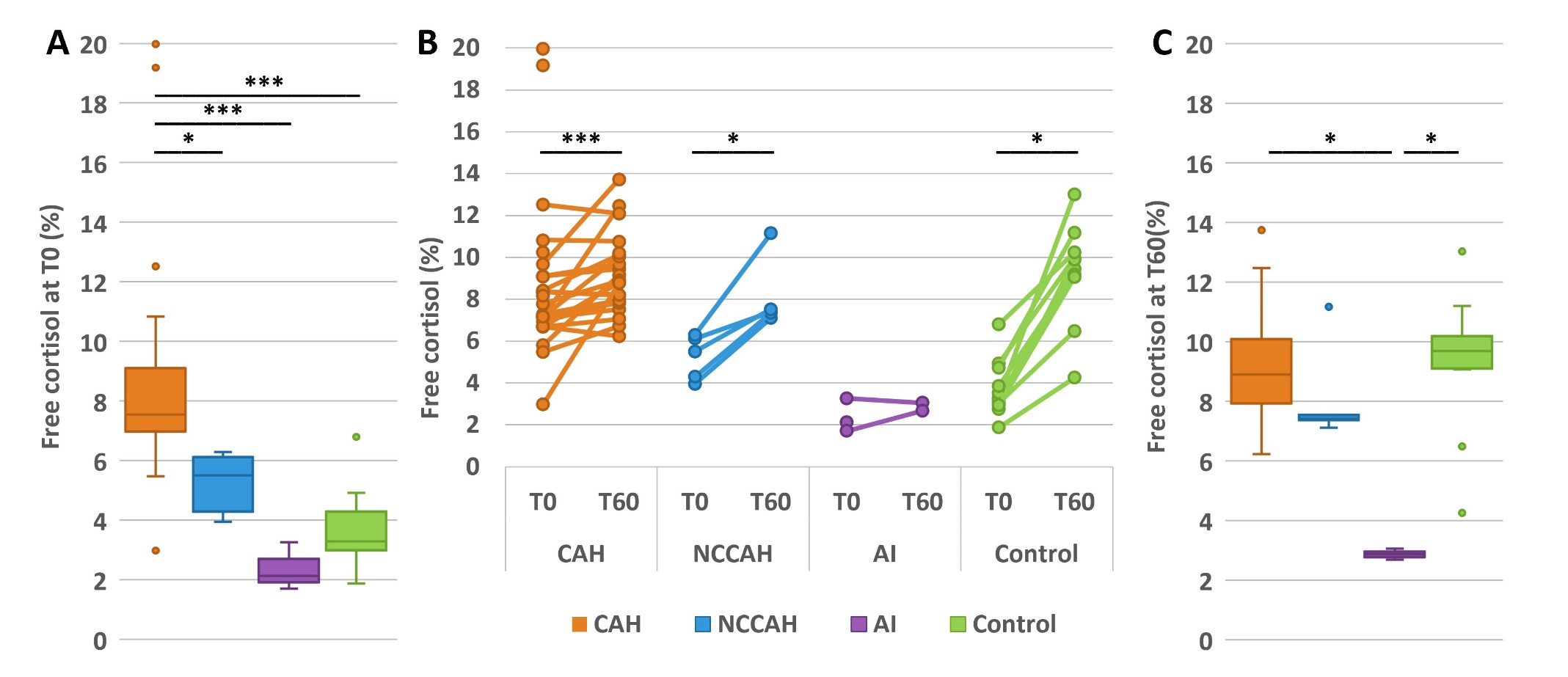

***Figure S2*** *Percentage of free cortisol before (T0) and after (T60) administration of Synacthen® in patients with classic congenital adrenal hyperplasia (CAH), non-classic congenital adrenal hyperplasia (NCCAH), adrenal insufficiency but not CAH (AI), and controls. (A) Free cortisol percentage at T0; (B) Change in free cortisol percentages between T0 and T60; (C) Free cortisol percentage at T60. *p≤0.05, ** p≤0.01, ***p≤0.001.*
